# Role of *BAG3* protein interactions in cardiomyopathies

**DOI:** 10.1101/2024.09.09.24313318

**Authors:** Hui-Qi Qu, JuFang Wang, Alexandre Rosa-Campos, Hakon Hakonarson, Arthur M. Feldman

## Abstract

**Background:** Bcl-2-associated athanogene 3 (BAG3) plays an important function in cellular protein quality control (PQC) maintaining proteome stability. Mutations in the BAG3 gene result in cardiomyopathies. Due to its roles in cardiomyopathies and the complexity of BAG3-protein interactions, it is important to understand these protein interactions given the importance of the multifunctional cochaperone BAG3 in cardiomyocytes, using an in vitro cardiomyocyte model.

**Methods:** The experimental assay was done using high pressure liquid chromatography coupled to tandem mass spectrometry (LC-MS/MS) in the human AC16 cardiomyocyte cell line with the BioID technology.

**Results:** Proteins with BAG3-interaction were identified in all the 28 hallmark gene sets enriched in idiopathic cardiomyopathies and/or ischemic disease. Among the 24 hallmark gene sets enriched in both idiopathic cardiomyopathies and ischemic disease, 15 gene sets had at least 3 proteins with BAG3-interaction.

**Conclusions:** This study highlights BAG3 protein interactions, unveiling the key gene sets affected in cardiomyopathies, which help to explain the molecular mechanisms of the cardioprotective effects of BAG3. In addition, this study also highlighted the complexity of proteins with BAG3 interactions, implying unwanted effects of BAG3.

## Introduction

BAG cochaperone 3 (BAG3), also known as Bcl-2-assciated athanogene 3, is a multi-functional protein, which is expressed ubiquitously in animals and homologs have been reported in plants^1^. BAG3 was first recognized for its ability to bind to Bcl2 with subsequent inhibition of apoptosis while other studies have found that it supports a diverse array of cellular functions including autophagy^2^, excitation-contraction coupling, mitochondrial function^3^, and the integrity of the sarcomere^4^. BAG3 has been recognized as playing a critical role in cellular protein quality control (PQC) to maintain the health of the proteome^5^. Mutations in the *BAG3* gene result in both dilated cardiomyopathy (DCM)^6^ and peripartum cardiomyopathy (PPCM)^7^ in a dominant inheritance model. Critical roles of *BAG3* in cardiomyocytes involves the maintenance of mitochondrial homeostasis that is mediated by both heat shock protein 70 (Hsp70)^8^ and the small heat shock proteins HspB6 and HspB8 (ref). BAG3 also has anti-apoptosis activity by binding Bcl-2^9^. BAG3 has 4 protein binding domains, including (1) 1 WW (Trp-Trp) domain, binding with proteins in signal transduction processes^10^, e.g. the PDZ domain containing guanine nucleotide exchange factor 2 (PDZGEF2) to promote cell adhesion^11^; (2) 2 IPV (Ile-Pro-Val) motifs, binding with small heat shock proteins (sHsps) HspB6/HspB8^12^; (3) 1 proline-rich repeat (PXXP) region, binding with SH3 (Src homology 3) motifs, e.g., in phospholipase C-γ (PLC-γ) that also serves as an attachment site for the dineen motor transport of misfolded proteins to the peri-nuclear aggresomes ^13,14^; and (4) 1 BAG domain, binding with Hsp70 and Bcl-2^12-14^.

While it is true that BAG3 is recognized for its multifunctionality and its role has been extensively studied, there is still much to uncover about its complex interactions and functions, especially in the context of different diseases. For instance, while the association of BAG3 with both cardiomyopathy and Parkinson’s disease (PD) has been reported, the genetic associations are in the opposite directions, i.e., the risk allele in DCM is protective against PD. The DCM-associated SNP rs2234962^15,16^ is in tight linkage disequilibrium with the PD-associated SNP rs72840788 (r^2^ = 1 in European populations) ^17,18^. We advocate for a greater focus on the intricate BAG3 interaction network, as a reductionist approach may fail to capture the subtleties and multifaceted nature of BAG3’s role. Due to the complexity of BAG3-protein interactions, it is useful to gain a better understanding of the specific proteins with which BAG3 participates in its activity as a multifunctional cochaperone by using an *in vitro* cell model to better understand BAG3 binding with other cellular proteins. For this purpose, we performed a proteomics study using the BioID proximity-dependent biotinylation method to identify proteins that interact with BAG3, particularly those from the gene sets with expression levels correlated with cardiomyopathies. BioID is a unique technology to screen for protein interactions in living cells^19^. In addition to direct protein interactions, BioID is able to identify weak or transient interactions, as well as proteins in close proximity. This discovery experiment was carried out to identify potentially novel proteins that bind to or interact with BAG3.

## Research Design and Methods

### Gene expression data

Gene expression data in tissue samples from left ventricular myocardium in two disease conditions, idiopathic and ischemic cardiomyopathies, were made available by Hannenhalli et al^20^. The study included 16 controls, 86 idiopathic, and 108 ischemic cardiomyopathies. The heart tissue was snap-frozen at time of cardiac transplantation. The gene expression assay was based on data generated using the Affymetrix Human Genome U133A Array. The data analysis was done by the GEO2R (https://www.ncbi.nlm.nih.gov/geo/info/geo2r.html). P values were adjusted by the Benjamini and Hochberg false discovery rate method. The data are publicly available at the NCBI Gene Expression Omnibus (GEO) database (https://www.ncbi.nlm.nih.gov/geo/query/acc.cgi?acc=GSE5406).

Gene Set Enrichment Analysis (GSEA) was done by the GSEA v4.3.2 software (Broad Institute of MIT and Harvard, MA) based on the Molecular Signatures Database (MSigDB)^21^ hallmark gene set collection^22^. FDR corrected p-values <0.05 were considered statistically significant.

### Cell experimental assay

The cell model used in this study was human AC16 cardiomyocyte cell line derived from adult human ventricular heart tissues (SCC109, Sigma-Aldrich). The cells were treated and analyzed based on five different conditions: (1) BAG3_Biotin: Cells transfected by plasmid expressing BioID-BAG3 for 48 hours, then get Biotin treatment for 16 hours; (2) BioID: Cells transfected by plasmid expressing BioID vector only for 48 hours, then get Biotin treatment for 16 hours; (3) BAG3 (to correct protein over-expression by BAG3): Cells transfected by plasmid expressing BioID-BAG3, without followed Biotin treatment; (4) AC16_Biotin (to correct protein-Biotin interaction): Cells get Biotin treatment for 16 hours; (5) AC16: Cells only. Three replicates were performed simultaneously for each treatment condition. Details of *BAG3* construct delivered using adeno-associated virus (AAV) vector has been described in our previous study^23^. The BioID experimental assay was done using high pressure liquid chromatography coupled to tandem mass spectrometry (LC-MS/MS) previously described through a collaboration with the Proteomics Facility, Sanford-Burnham-Presby Medical Discovery Institute, La Jolla, CA^24^.

### Data analysis of BAG3-Protein interactions

The BioID data were analyzed using MSstats package from R Bioconductor^25^. By comparing the groups of BAG3_Biotin *vs*. BioID, all proteins with adjusted P-value<0.05 by the Benjamini and Hochberg false discovery rate method were identified. The effect sizes of BAG3-protein interactions were corrected by [log2FC(BAG3_Biotin vs BioID)]-[log2FC(AC16_Biotin vs AC16)]-[log2FC(BAG3 vs BioID)], i.e. BAG3-protein interactions being corrected for biotin-protein interactions and BAG3-increased protein levels.

## Results

### Gene expression in idiopathic and ischemic cardiomyopathies

Among the 50 hallmark gene sets, 26 showed significance in idiopathic CM, and 26 showed significances in ischemic CM (Table 1, Fig.1). From these gene sets, 24 gene sets were significant in both idiopathic and ischemic CMs, while the other 4 gene sets had significances in only idiopathic or ischemic CMs. For the later 4 gene sets, the same direction trend of enrichment was observed in the other type of CM. In particular, *BAG3* levels were decreased in both idiopathic (fold change=0.646, adjusted P-value=9.60E-06) and ischemic (fold change =0.633, adjusted P-value=9.77E-06) cardiomyopathies.

**Figure 1.**
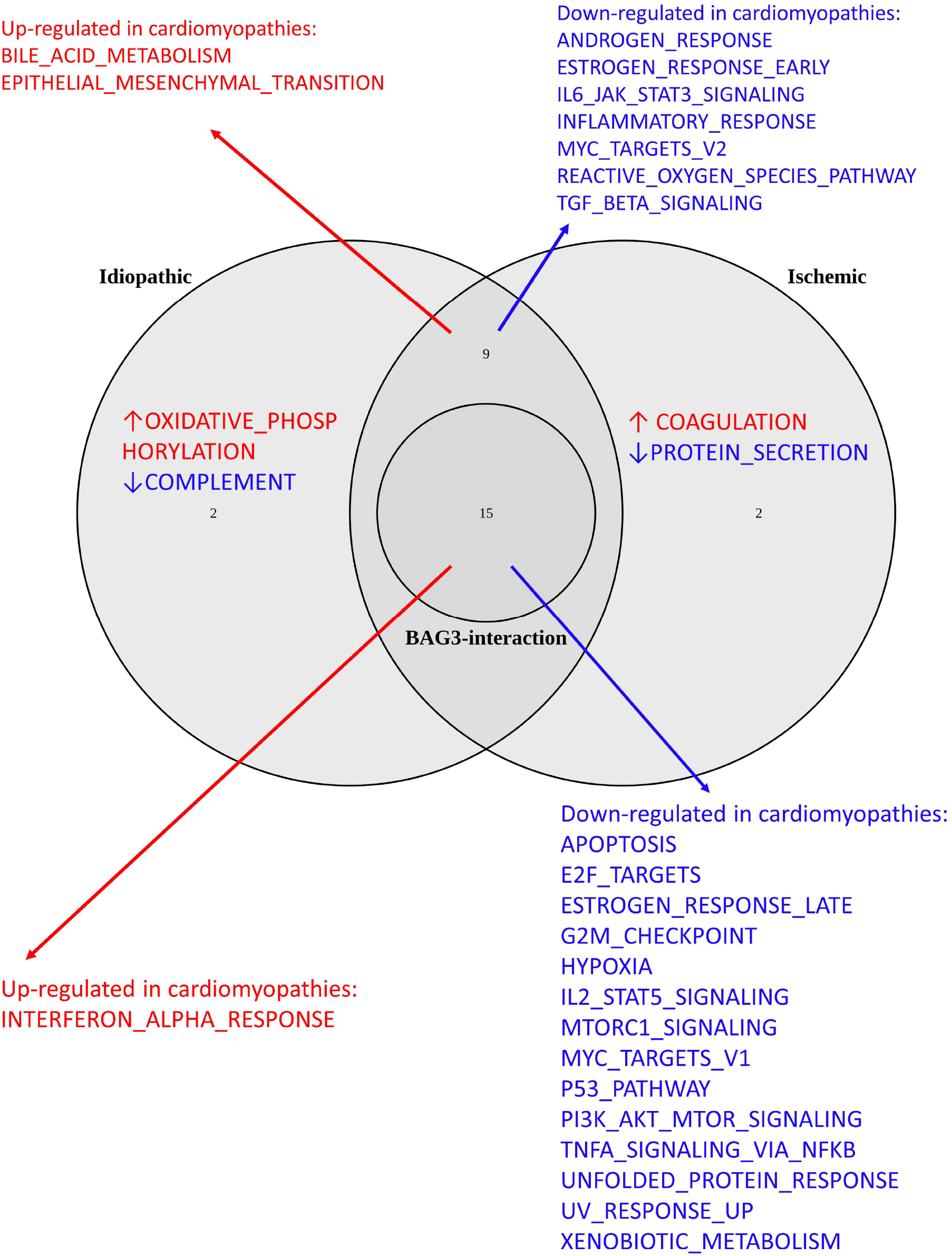
Venn plot of Hallmark gene sets enriched in cardiomyopathies and those with BAG3 interactions.

**Table 1.**
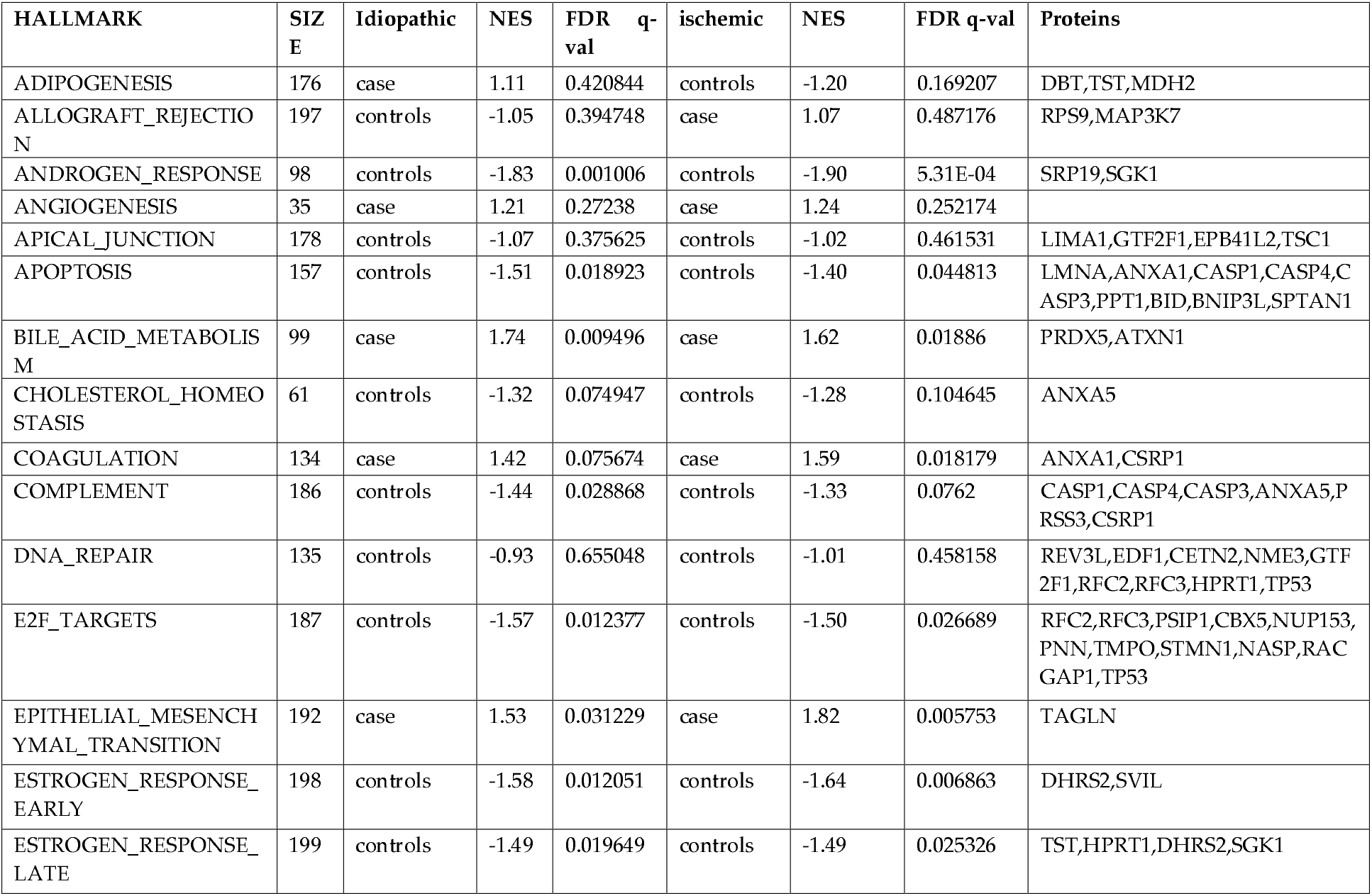
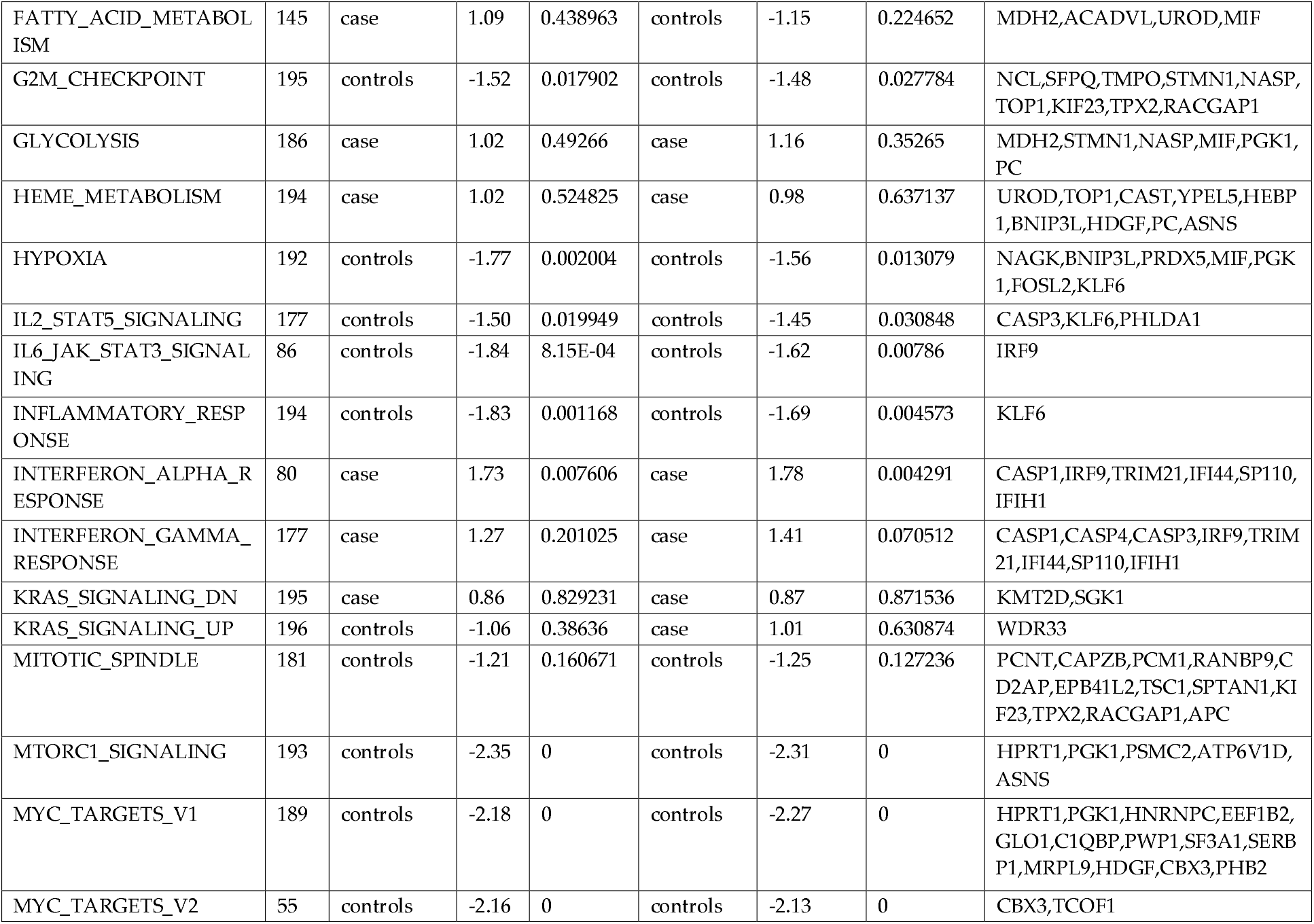
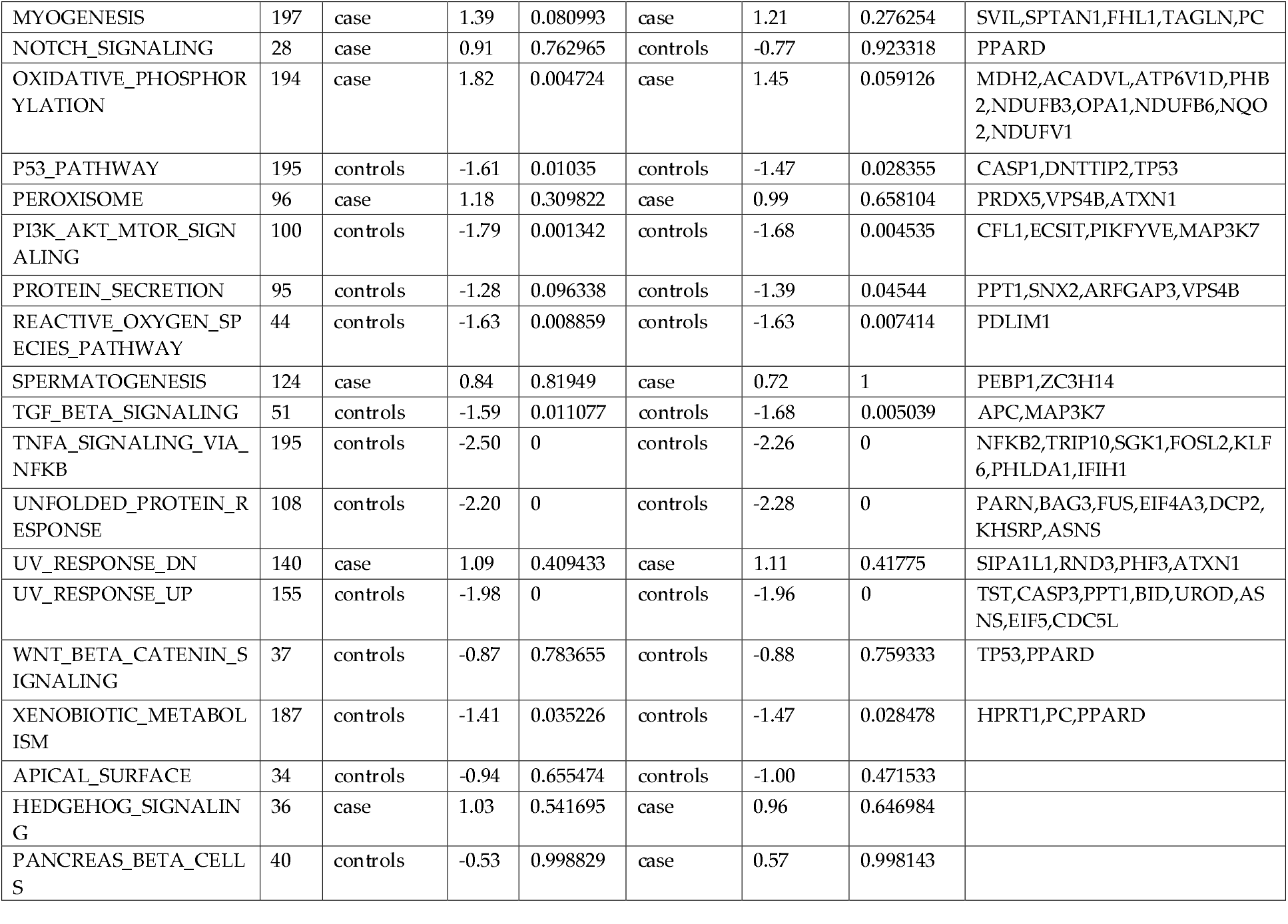
50 hallmark gene sets in idiopathic and ischemic cardiomyopathies.

| HALLMARK | SIZE | Idiopathic | NES | FDR q-val | ischemic | NES | FDR q-val | Proteins |
| --- | --- | --- | --- | --- | --- | --- | --- | --- |
| ADIPOGENESIS | 176 | case | 1.11 | 0.420844 | controls | -1.20 | 0.169207 | DBT,TST,MDH2 |
| ALLOGRAFT_REJECTION | 197 | controls | -1.05 | 0.394748 | case | 1.07 | 0.487176 | RPS9,MAP3K7 |
| ANDROGEN_RESPONSE | 98 | controls | -1.83 | 0.001006 | controls | -1.90 | 5.31E-04 | SRP19,SGK1 |
| ANGIOGENESIS | 35 | case | 1.21 | 0.27238 | case | 1.24 | 0.252174 |  |
| APICAL_JUNCTION | 178 | controls | -1.07 | 0.375625 | controls | -1.02 | 0.461531 | LIMA1,GTF2F1,EPB41L2,TSC1 |
| APOPTOSIS | 157 | controls | -1.51 | 0.018923 | controls | -1.40 | 0.044813 | LMNA,ANXA1,CASP1,CASP4,CASP3,PPT1,BID,BNIP3L,SPTAN1 |
| BILE_ACID_METABOLISM | 99 | case | 1.74 | 0.009496 | case | 1.62 | 0.01886 | PRDX5,ATXN1 |
| CHOLESTEROL_HOMEOSTASIS | 61 | controls | -1.32 | 0.074947 | controls | -1.28 | 0.104645 | ANXA5 |
| COAGULATION | 134 | case | 1.42 | 0.075674 | case | 1.59 | 0.018179 | ANXA1,CSRP1 |
| COMPLEMENT | 186 | controls | -1.44 | 0.028868 | controls | -1.33 | 0.0762 | CASP1,CASP4,CASP3,ANXA5,PRSS3,CSRP1 |
| DNA_REPAIR | 135 | controls | -0.93 | 0.655048 | controls | -1.01 | 0.458158 | REV3L,EDF1,CETN2,NME3,GTF2F1,RFC2,RFC3,HPRT1,TP53 |
| E2F_TARGETS | 187 | controls | -1.57 | 0.012377 | controls | -1.50 | 0.026689 | RFC2,RFC3,PSIP1,CBX5,NUP153,PNN,TMPO,STMN1,NASP,RACGAP1,TP53 |
| EPITHELIAL_MESENCHYMAL_TRANSITION | 192 | case | 1.53 | 0.031229 | case | 1.82 | 0.005753 | TAGLN |
| ESTROGEN_RESPONSE_EARLY | 198 | controls | -1.58 | 0.012051 | controls | -1.64 | 0.006863 | DHRS2,SVIL |
| ESTROGEN_RESPONSE_LATE | 199 | controls | -1.49 | 0.019649 | controls | -1.49 | 0.025326 | TST,HPRT1,DHRS2,SGK1 |
| FATTY_ACID_METABOLISM | 145 | case | 1.09 | 0.438963 | controls | -1.15 | 0.224652 | MDH2,ACADVL,UROD,MIF |
| G2M_CHECKPOINT | 195 | controls | -1.52 | 0.017902 | controls | -1.48 | 0.027784 | NCL,SFPQ,TMPO,STMN1,NASP, TOP1,KIF23,TPX2,RACGAP1 |
| GLYCOLYSIS | 186 | case | 1.02 | 0.49266 | case | 1.16 | 0.35265 | MDH2,STMN1,NASP,MIF,PGK1, PC |
| HEME_METABOLISM | 194 | case | 1.02 | 0.524825 | case | 0.98 | 0.637137 | UROD, TOP1, CAST, YPEL5, HEBP 1, BNIP3L, HDGF, PC, ASNS |
| HYPOXIA | 192 | controls | -1.77 | 0.002004 | controls | -1.56 | 0.013079 | NAGK, BNIP3L, PRDX5, MIF, PGK 1, FOSL2, KLF6 |
| IL2_STAT5_SIGNALING | 177 | controls | -1.50 | 0.019949 | controls | -1.45 | 0.030848 | CASP3, KLF6, PHLDA1 |
| IL6_JAK_STAT3_SIGNALING | 86 | controls | -1.84 | 8.15E-04 | controls | -1.62 | 0.00786 | IRF9 |
| INFLAMMATORY_RESPONSE | 194 | controls | -1.83 | 0.001168 | controls | -1.69 | 0.004573 | KLF6 |
| INTERFERON_ALPHA_RESPONSE | 80 | case | 1.73 | 0.007606 | case | 1.78 | 0.004291 | CASP1, IRF9, TRIM21, IFI44, SP110, IFIH1 |
| INTERFERON_GAMMA_RESPONSE | 177 | case | 1.27 | 0.201025 | case | 1.41 | 0.070512 | CASP1, CASP4, CASP3, IRF9, TRIM 21, IFI44, SP110, IFIH1 |
| KRAS_SIGNALING_DN | 195 | case | 0.86 | 0.829231 | case | 0.87 | 0.871536 | KMT2D, SGK1 |
| KRAS_SIGNALING_UP | 196 | controls | -1.06 | 0.38636 | case | 1.01 | 0.630874 | WDR33 |
| MITOTIC_SPINDLE | 181 | controls | -1.21 | 0.160671 | controls | -1.25 | 0.127236 | PCNT, CAPZB, PCM1, RANBP9, C D2AP, EPB41L2, TSC1, SPTAN1, KIF23, TPX2, RACGAP1, APC |
| MTORC1_SIGNALING | 193 | controls | -2.35 | 0 | controls | -2.31 | 0 | HPRT1, PGK1, PSMC2, ATP6V1D, ASNS |
| MYC_TARGETS_V1 | 189 | controls | -2.18 | 0 | controls | -2.27 | 0 | HPRT1, PGK1, HNRNPC, EEF1B2, GLO1, C1QBP, PWP1, SF3A1, SERB P1, MRPL9, HDGF, CBX3, PHB2 |
| MYC_TARGETS_V2 | 55 | controls | -2.16 | 0 | controls | -2.13 | 0 | CBX3, TCOF1 |
| MYOGENESIS | 197 | case | 1.39 | 0.080993 | case | 1.21 | 0.276254 | SVIL,SPTAN1,FHL1,TAGLN,PC |
| NOTCH_SIGNALING | 28 | case | 0.91 | 0.762965 | controls | -0.77 | 0.923318 | PPARD |
| OXIDATIVE_PHOSPHORYLATION | 194 | case | 1.82 | 0.004724 | case | 1.45 | 0.059126 | MDH2,ACADVL,ATP6V1D,PHB2,NDUFB3,OPA1,NDUFB6,NQO2,NDUFV1 |
| P53_PATHWAY | 195 | controls | -1.61 | 0.01035 | controls | -1.47 | 0.028355 | CASP1,DNTTIP2,TP53 |
| PEROXISOME | 96 | case | 1.18 | 0.309822 | case | 0.99 | 0.658104 | PRDX5,VPS4B,ATXN1 |
| PI3K_AKT_MTOR_SIGNALING | 100 | controls | -1.79 | 0.001342 | controls | -1.68 | 0.004535 | CFL1,ECSIT,PIKFYVE,MAP3K7 |
| PROTEIN_SECRETION | 95 | controls | -1.28 | 0.096338 | controls | -1.39 | 0.04544 | PPT1,SNX2,ARFGAP3,VPS4B |
| REACTIVE_OXYGEN_SPECIES_PATHWAY | 44 | controls | -1.63 | 0.008859 | controls | -1.63 | 0.007414 | PDLIM1 |
| SPERMATOGENESIS | 124 | case | 0.84 | 0.81949 | case | 0.72 | 1 | PEBP1,ZC3H14 |
| TGF_BETA_SIGNALING | 51 | controls | -1.59 | 0.011077 | controls | -1.68 | 0.005039 | APC,MAP3K7 |
| TNFA_SIGNALING_VIA_NFKB | 195 | controls | -2.50 | 0 | controls | -2.26 | 0 | NFKB2,TRIP10,SGK1,FOSL2,KLF6,PHLDA1,IFIH1 |
| UNFOLDED_PROTEIN_RESPONSE | 108 | controls | -2.20 | 0 | controls | -2.28 | 0 | PARN,BAG3,FUS,EIF4A3,DCP2,KHSRP,ASNS |
| UV_RESPONSE_DN | 140 | case | 1.09 | 0.409433 | case | 1.11 | 0.41775 | SIPA1L1,RND3,PHF3,ATXN1 |
| UV_RESPONSE_UP | 155 | controls | -1.98 | 0 | controls | -1.96 | 0 | TST,CASP3,PPT1,BID,UROD,ASNS,EIF5,CDC5L |
| WNT_BETA_CATENIN_SIGNALING | 37 | controls | -0.87 | 0.783655 | controls | -0.88 | 0.759333 | TP53,PPARD |
| XENOBIOTIC_METABOLISM | 187 | controls | -1.41 | 0.035226 | controls | -1.47 | 0.028478 | HPRT1,PC,PPARD |
| APICAL_SURFACE | 34 | controls | -0.94 | 0.655474 | controls | -1.00 | 0.471533 |  |
| HEDGEHOG_SIGNALING | 36 | case | 1.03 | 0.541695 | case | 0.96 | 0.646984 |  |
| PANCREAS_BETA_CELLS | 40 | controls | -0.53 | 0.998829 | case | 0.57 | 0.998143 |  |

### BAG3-Protein interactions

387 proteins were identified with significant adjusted P-values and positive BAG3-protein interactions (Supplementary Table 1). Proteins with BAG3-interaction were identified in each of 28 hallmark gene sets enriched in the idiopathic and/or ischemic cardiomyopathies. Among the 24 hallmark gene sets enriched in both idiopathic and ischemic cardiomyopathies, 15 gene sets had at least 3 proteins with BAG3-interaction (Table 2, Fig.1).

**Table 2.** 15 HALLMARK gene sets with at least 3 BAG3-interacted proteins.

| Gene set | Note | BAG3-interacted Proteins |
| --- | --- | --- |
| **Down-regulated in cardiomyopathies** |  |  |
| MYC_TARGETS_V1 | Signal DNA damage-induced apoptosis<sup>37</sup> | HPRT1,PGK1,HNRNPC,EEF1B2,GL O1,C1QBP,PWP1,SF3A1,SERBP1,MR PL9,HDGF,CBX3,PHB2 |
| E2F_TARGETS | Integrate cell cycle progression with DNA repair, replication, and G2/M checkpoints<sup>34</sup> | RFC2,RFC3,PSIP1,CBX5,NUP153,PN N,TMPO,STMN1,NASP,RACGAP1, TP53 |
| G2M_CHECKPOINT | Prevent DNA damaged cells from entering mitosis<sup>39</sup> | NCL,SFPQ,TMPO,STMN1,NASP,TO P1,KIF23,TPX2,RACGAP1 |
| P53_PATHWAY | Promote cell cycle arrest to allow DNA repair or apoptosis<sup>38</sup> | CASP1,DNTTIP2,TP53 |
| APOPTOSIS | Cardiomyocyte apoptosis-interruptus<sup>48</sup> | LMNA,ANXA1,CASP1,CASP4,CAS P3,PPT1,BID,BNIP3L,SPTAN1 |
| HYPOXIA | Alter myocardial gene expression and induce cardiomyocyte apoptosis<sup>52</sup> | NAGK,BNIP3L,PRDX5,MIF,PGK1,F OSL2,KLF6 |
| XENOBIOTIC_METABOLISM | Oxidative stress with excessive production of reactive oxygen species (ROS)<sup>55</sup> | HPRT1,PC,PPARD |
| UV_RESPONSE_UP | DNA damage repair<sup>53</sup> | TST,CASP3,PPT1,BID,UROD,ASNS, EIF5,CDC5L |
| UNFOLDED_PROTEIN_RESPONSE | Decrease global protein synthesis, and increase refolding or degradation of misfolded proteins<sup>56</sup> | PARN,BAG3,FUS,EIF4A3,DCP2,KH SRP,ASNS |
| TNFA_SIGNALING_VIA_NFKB | Cardioprotective in hypoxic or ischemic myocardial injury<sup>64</sup> | NFKB2,TRIP10,SGK1,FOSL2,KLF6,P HLDA1,IFIH1 |
| MTORC1_SIGNALING | Regulate cell growth and metabolism<sup>66</sup> and mediate adaptive cardiac hypertrophy<sup>67</sup> | HPRT1,PGK1,PSMC2,ATP6V1D,AS NS |
| ESTROGEN_RESPONSE_LATE | Prevent apoptosis and necrosis of cardiac and endothelial cells<sup>69</sup> | TST,HPRT1,DHRS2,SGK1 |
| PI3K_AKT_MTOR_SIGNALING | Lead to cell proliferation<sup>70</sup> and inhibit cardiomyocyte apoptosis<sup>71</sup> | CFL1,ECSIT,PIKFYVE,MAP3K7 |
| IL2_STAT5_SIGNALING | Modulate CD4+ Th cell differentiation<sup>72</sup> and upregulate the expression of c-Myc, BCL-2, and BCL-x<sup>73</sup> | CASP3,KLF6,PHLDA1 |
| **Up-regulated in cardiomyopathies** |  |  |
| INTERFERON_ALPHA_RESPONSE | Interferon inhibits cardiac cell function *in vitro*<sup>74</sup> and interferon treatment has been shown of cardiotoxicity<sup>75</sup>. | CASP1,IRF9,TRIM21,IFI44,SP110,IFIH1 |

## Discussion

This study identified 387 proteins with direct or indirect BAG3 interactions. Among these proteins, heat shock 70 kDa protein 6 (HSPA6)^26^ and Hsp70-binding protein 1 (HSPBP1)^27^ encode a Hsp70 protein and regulate Hsp70 function, respectively. Bcl-2-associated transcription factor 1 (BCLAF1) actives the p53 pathway and induces apoptosis^28^, and may contribute to myocardial reperfusion injury^29^. BCL2/adenovirus E1B 19 kDa protein-interacting protein 3-like (BNIP3L) activates the ER and mitochondrial cell death pathways, and induces cardiomyopathy^30^. In addition, we observed 15 gene sets with at least 3 BAG3-interacted proteins further supporting the important role of BAG3 in the biology of the heart (Table 2). In a previous study, Chen et al. identified 382 BAG3-interacting proteins in cancer cell lines using stable isotope labeling with amino acids in cell culture (SILAC) combined with mass spectrometry (MS)^31^. Of the 387 proteins identified in our study, 22 overlapped with those reported by Chen et al. Given the estimated 20,000 protein-coding genes in the human genome, this overlap is highly statistically significant and unlikely to occur by chance alone (P=3.69E-07). Furthermore, many of the 360 proteins identified by Chen et al. but not replicated in our study are closely associated with cancer-related processes, including glutathione metabolism, disruptions in protein catabolism, protein folding, ubiquitin-dependent degradation, apoptosis evasion, post-translational modifications, and the regulation of protein complex assembly. Notably, 365 of the proteins identified in our study have not been reported by Chen et al.

### Proteins with roles in cell cycle

Cardiomyocytes are taken as terminally differentiated and don’t proliferate after birth^32^. In a recent study, a proliferative burst in preadolescent mice was observed^33^. However, further experimental validation is needed, as this finding has not yet been replicated by other researchers. If the same phenomenon is true in human, the highlighted cell cycle proteins in this study may imply for early intervention for cardiomyopathies, particularly for a BAG3-based therapy. Adult cardiomyocytes also exhibit a dynamic range of cell cycle activity under various physiological and pathological conditions, e.g. in pathologic myocardial hypertrophy^34^.

BAG3-interaction was identified in 31 gene proteins from four important gene sets in cell cycle, including MYC_TARGETS_V1, E2F_TARGETS, G2M_CHECKPOINT, and P53_PATHWAY. These four gene sets were down-regulated in both idiopathic and ischemic cardiomyopathies. MYC, E2F and p53 are important regulators of cell-cycle progression^35^. We observed BAG3 interaction of 13 proteins involving MYC signaling, including target variant 1 proteins, 11 proteins in E2F signaling molecules, 9 proteins in cell cycle G2/M checkpoint, and 3 proteins in the p53 pathway. The MYC_TARGETS_V1 genes are regulated by MYC^22^, and involved in cell-cycle progression and cell proliferation^36^. c-Myc plays an essential role in signaling DNA damage-induced apoptosis through the control of the p53 tumor suppressor protein^37^. Activation of p53 pathway promotes cell cycle arrest to allow DNA repair or apoptosis for cells with serious DNA damage^38^. The G2/M checkpoints prevent DNA damaged cells from entering mitosis^39^. Acting through the E2F targets, E2F integrates cell cycle progression with DNA repair, replication, and G2/M checkpoints^34^.

The proteins with BAG3-interaction in these gene sets also harbor opportunities for small molecular therapies. Deficiency of the MYC target gene phosphoglycerate kinase 1 (*PGK1*) from genetic mutations may cause myopathy^40^. The protein encoded by *PGK1* catalyzes the production if adenosine 5’-triphosphate (ATP) and its activity is regulated by ATP^41^. Biallelic deficiency of the MYC target gene complement component 1 Q subcomponent-binding protein, mitochondrial (*C1QBP*) cause mitochondrial respiratory-chain deficiencies and severe cardiomyopathy^42^. Copper supplementation has been shown to up-regulate the function of the *C1QBP* protein and may improve cardiac function^43^.

### 2 Interactions with proteins involved in apoptosis and cell damage responses

In addition to the cell cycle proteins, BAG3 interacts with proteins in a number of gene sets downregulated in cardiomyopathies, including HALLMARK_APOPTOSIS, HALLMARK_HYPOXIA, HALLMARK_UV_RESPONSE_UP, HALLMARK_XENOBIOTIC_METABOLISM, HALLMARK_UNFOLDED_PROTEIN_RESPONSE, HALLMARK_PROTEIN_SECRETION, HALLMARK_COMPLEMENT, HALLMARK_TNFA_SIGNALING_VIA_NFKB, HALLMARK_MTORC1_SIGNALING, HALLMARK_ESTROGEN_RESPONSE_LATE, HALLMARK_PI3K_AKT_MTOR_SIGNALING, and

HALLMARK_IL2_STAT5_SIGNALING. BAG3 interactions with apoptosis process, responses to DNA damage and oxidative stress, and cell repairs were highlighted.

It has been suggested that cardiomyocyte apoptosis may contribute to myocardial reperfusion injury^44,45^ and cardiomyopathies ^46,47^. Currently, there is controversial evidence regarding the involvement of apoptosis in cardiomyopathies. Narula et al. ^48^ described an interruption in the apoptosis cascade, including nuclear fragmentation and condensation in cardiomyocytes, terming it ‘apoptosis-interruptus.’ Conversely, our studies and others suggest that apoptosis plays a critical role in heart failure^49^.

The gene expression data of cardiomyopathies show that the apoptosis pathway is down-regulated in both idiopathic and ischemic cardiomyopathies. Cardiomyocytes with interrupted apoptosis may undergo necrosis. BAG3 binds to the Bcl-2 Homology 4 (BH4) domain of Bcl-2 to inhibit mitochondrial-dependent (intrinsic) apoptosis^9^, thus protect from cell death. Nine proteins in the APOPTOSIS gene set were identified of BAG3 interaction in this study, including 3 caspases, including CASP1, CASP3, and CASP4.

Hypoxia significantly alters myocardial gene expression, mediated by hypoxia inducible factor 1 subunit alpha (HIF1A)^50^. Hypoxia has been shown to induce dilated cardiomyopathy in chick embryos^51^. Overexpression of BAG3 has been shown to attenuate hypoxia-induced cardiomyocyte apoptosis^52^. DNA damage induces apoptosis and cardiomyopathy^53^, whereas HALLMARK_UV_RESPONSE_UP genes (i.e,. up-regulated genes in response to UV), play major roles in DNA damage repair^54^. Moreover, Xenobiotic toxicity causes cardiomyopathy through oxidative stress^55^ resuling in dysregulated gene expression.

The unfolded protein response (UPR) decreases global protein synthesis, increases endoplasmic reticulum (ER)-associated degradation of misfolded proteins, and activates protein-folding in ER^56^. BAG3 serves as a cochaperone with Hsp70 and is involved in a wide range of protein folding processes, including refolding of misfolded^57^. The protein secretion pathway acts on protein folding, post-translational modifications (PTMs), and protein trafficking^58^, and is important in cardiac repair^59^.

TNF-α^60^and complement activation^61^ have been suggested as contributing factors to myocardial reperfusion injury and cardiomyopathy. However, complement activation contributes to tissue repair^62^. the genes encoding components of the complement system aredownregulated in idiopathic and ischemic cardiomyopathies, which is in consistent with the recognition that long-term effects of complement inhibitors may be detrimental^63^. While NF-κB signaling may be cardioprotective in hypoxic or ischemic myocardial injury^64^, it has also been shown to mediate chronic inflammation^65^. The mTORC1 signaling regulates cell growth and metabolism^66^ and mediates adaptive cardiac hypertrophy^67^. Its inhibition increases overall protein degradation by the ubiquitin proteasome system^68^, and may attenuate cardiac remodeling and heart failure^67^. Late estrogen response pathway may prevent apoptosis and necrosis of cardiac and endothelial cells^69^. Genes up-regulated by activation of the PI3K/AKT/mTOR pathway, include a major intracellular network that leads to cell proliferation^70^ and inhibits cardiomyocyte apoptosis^71^. IL2_JAK_STAT5_SIGNALING genes are up-regulated by STAT5 in response to IL2 stimulation and modulate CD4+ Th cell differentiation^72^, and upregulate the expression of c-Myc, BCL-2, and BCL-x further exacerbating the inflammatory response^73^.

### 3 Interactions with proteins upregulated in cardiomyopathies

In contrast to the above gene sets, the HALLMARK_INTERFERON_ALPHA_RESPONSE and HALLMARK_OXIDATIVE_PHOSPHORYLATION gene sets are both up-regulated in cardiomyopathies. Interferon inhibits cardiac cell function *in vitro*^74^ and interferon treatment has been shown to produce cardiotoxicity^75^. The heart is in high demand of energy by oxidative phosphorylation, where defective oxidative phosphorylation may cause cardiomyopathy^76^. Oxidative phosphorylation genes are upregulated in cardiomyopathies, which may cause over-production of reactive oxygen species (ROS)^77^. BAG3 interactions with these upregulated genes in cardiomyopathies is interesting as this could ameliorate the cardioprotective effects of BAG3. Adjunct therapy targeting these proteins may thus improve the therapeutic effects of BAG3. The NADH dehydrogenases, i.e., NDUFB3 (NADH dehydrogenase 1 beta subcomplex subunit 3), NDUFB6 (NADH dehydrogenase 1 beta subcomplex subunit 6), and NDUFV1 (NADH dehydrogenase flavoprotein 1, mitochondrial), can be inhibited by metformin. The potential heart protective effects by metformin are gaining more research attention^78^.

## Conclusion

This study utilized the relatively new technique of BioID to identify interactions between BAG3 and various gene sets. The goal was to uncover unique proteins and protein pathways that might contribute to disease when BAG3 levels are under-expressed. Such under-expression can occur due to loss-of-function mutations caused by truncations, deletions, or unique mutations secondary to single nucleotide variants. These variants may change an amino acid, insert an amino acid, or alter the reading frame, resulting in truncation. Not unexpectedly, we identified a plethora of associated proteins. In view of the small size of BAG3, the limited number of binding sites, and the somewhat focused activities of the protein, the number of intersecting proteins identified by this new technique represents an overabundance of detected proteins. However, the fact that many of the observed proteins have been associated with BAG3 in various animal studies raises the larger question of how a single protein might interact with a large number of target proteins. The fact that virtually every known protein interaction with BAG3 has been detected by the BioID assay suggests that enhancing the assay’s sensitivity might allow for a more nuanced approach to this question. Additionally, new means of discriminating interactome data, which remain proprietary, could be useful.

In conclusion, this study highlights the observed interactions of BAG3 with key gene sets that are affected in cardiomyopathies, thereby unveiling some of the molecular mechanisms involved with the cardioprotective effects of BAG3. In addition, this study also highlights the complexity of proteins with BAG3 interactions, implying unwanted effects of BAG3. Adjunct therapy to address unwanted effects of BAG3 may be indicated, such as the use of metformin to inhibit the consequences of NADH dehydrogenase activation. A limitation of this study is the AC16 cell model, which is an immortalized cell line with passages over numerous generations^79^, and may not represent the actual cardiomyocytes in human heart. Although AC16 cells are derived from human ventricles, they exhibit several key limitations: 1) they do not exhibit contractile activity, 2) they are highly proliferative, and 3) the sarcomere is not organized in AC16 cells. While AC16 cells differ from primary cardiomyocytes in these aspects, they remain a widely used *in vitro* model for studying human cardiomyocyte-related processes due to their human origin and ability to express cardiac-specific markers. The primary objective of our study was to investigate the complex BAG3 interaction network and identify its interacting proteins, so the aforementioned limitations do not significantly impact our findings. Further study using different cell models, in particular specialized cardiomyocyte models, will help to verify the BAG3 interactions, and enable investigations of the more focused phenotypic effects. Additionally, validation through co-immunoprecipitation (Co-IP) studies, although challenging to scale up and expensive for large-scale applications, will be important for confirming selected candidate proteins of interest in human heart tissues and human-induced pluripotent stem cell-derived cardiomyocytes (hIPSC-CMs) to corroborate our findings and enhance their relevance to the human heart.

## Data Availability

All data produced in the present study are available upon reasonable request to the authors.

## Supplementary Materials

Table S1: 387 proteins with BAG3-protein interactions.

## Author Contributions

Conceptualization, H.H. and A.M.F.; methodology, H.Q.Q.; formal anal-ysis, H.Q.Q.; investigation, H.Q.Q., J.W., and A.R.C.; resources, A.M.F.; data curation, H.Q.Q. and A.R.C.; writing—original draft preparation, H.Q.Q.; writing—review and editing, H.H. and A.M.F.; visualization, H.Q.Q.; supervision, H.H.; project administration, H.H. and A.M.F.; funding acquisition, H.H. and A.M.F. All authors have read and agreed to the published version of the manuscript.” Please turn to the CRediT taxonomy for the term explanation. Authorship must be limited to those who have contributed substantially to the work reported.

## Institutional Review Board Statement

This study was approved by the Institutional Review Board (IRB) of the Children’s Hospital of Philadelphia. Human participants and personal information are inaccessible to the research group. All human subjects or their legal guardians provided written informed consent.

## Informed Consent Statement

Not applicable.

## Data Availability Statement

Supporting data from this study can be obtained by emailing the corresponding author Dr. Hakon Hakonarson.

